# Reduced Intrinsic Neural Timescales in Schizophrenia along Posterior Parietal and Occipital Areas

**DOI:** 10.1101/2021.05.22.21257646

**Authors:** Lavinia Carmen Uscătescu, Sarah Said-Yürekli, Lisa Kronbichler, Renate Stelzig-Schöler, Brandy-Gale Pearce, Luise Antonia Reich, Stefanie Weber, Wolfgang Aichhorn, Martin Kronbichler

**Author notes:** Currently based at the Olin Neuropsychiatry Research Centre, Yale School of Medicine, Hartford, Connecticut, United States of America.

## Abstract

We computed intrinsic neural timescales (INT) based on resting state functional magnetic resonance imaging (rsfMRI) data of healthy controls (HC) and patients with schizophrenia spectrum disorder (SZ) from three independently collected samples. Five clusters showed decreased INT in SZ compared to HC in all three samples: Right occipital fusiform gyrus (rOFG), Left superior occipital gyrus (lSOG), Right superior occipital gyrus (rSOG), Left lateral occipital cortex (lLOC), and Right postcentral gyrus (rPG). In other words, it appears that sensory information in visual and posterior parietal areas is stored for reduced lengths of time in SZ compared to HC. We also found some evidence that symptom severity modulates INT of these areas in SZ.

## Introduction

Schizophrenia is a psychiatric disorder diagnosed in approximately 1% of the world’s population (Bhugra, 2005). It is characterized by negative (e.g. disorganized thoughts and language, attention and memory deficits) and positive (e.g. hallucinations and delusions) symptoms. Of high relevance to SZ pathology are the visual, auditory and sensorimotor areas. The dysconnectivity and disintegration of primary sensory areas have been proposed to underlie higher cognitive dysfunctions in SZ (Bordier et al., 2018; Ferri et al., 2017; Kaufmann et al., 2015) and have been shown to be predictive of disease severity (Guo et al., 2014; Javitt, 2015; Orliac et al., 2017; Zhang, Guo and Tian, 2019; Zhang et al., 2019). For example, increased connectivity between early and late visual areas has been linked to mood induction in a compensatory manner in SZ (Dyck et al., 2014). Furthermore, cognitive control deficits in SZ patients have been linked to hyper-connectivity within the auditory, sensorimotor and posterior parietal cortex (Mayer et al., 2015). The thalamus, involved in sensory gating deficits in SZ, has also been shown to be hyper-connected to sensorimotor areas (Xi et al., 2020), and its increased connectivity to the middle temporal gyrus has been positively related to the presence of hallucinations and delusions (Ferri et al., 2018). Finally, connectivity alterations in somatosensory areas have also shown to be good predictors of patient classification (Skåtun et al., 2016).

In later years, there has been a greater emphasis on characterising neuropsychiatric disorders in terms of trans-diagnostic, as opposed to categorical symptoms. One such symptom reflects sensory processing deficits (Hornix, Havekes and Kas, 2019; Harrison et al., 2019), which comprise responding to, processing and organizing sensory information (Miller et al., 2009). Sensory deficits have been found to characterize several neuropsychiatric disorders, such as SZ (Brown et al., 2002; Javitt and Freedman, 2015) and Autism Spectrum Disorders (ASD; Marco et al., 2011; Balasco, Provenzano and Bozzi, 2020). True to its trans-diagnostic potential, when comparing ASD and SZ directly, these deficits have been found to constitute a common feature of both disorders (Noel, Stevenson and Wallace, 2018; Zhou et al., 2018; Zhou et al., 2020).

Most neuroimaging research deals with analysing static relationships between functional neural components. However, this can only offer limited insight into brain health and disease, since brain activity is essentially dynamic. Consequently, a range of time series analyses directed at characterizing the dynamic changes in brain activity have been developed in recent years. Vince Calhoun et al. (2014) proposed the umbrella-term “chronnectome” to describe these dynamic brain processes. “Chronnectomic” approaches are diverse, can be applied to both M/EEG and fMRI time series, and have proven useful for distinguishing neurotypicals from, e.g., SZ (e.g. Miller, Yaesoubi, and Calhoun, 2014). One such chronnectomic approach is to assess how long information is stored in various neural areas. This duration is known as *temporal receptive field, intrinsic neural timescales (INT)*, or *temporal receptive window*.

A hierarchical organization of INT across the primate cortex has been initially noted based on spike count (Murray et al., 2014), with sensory areas displaying shorter INT compared to frontal ones. Human neuroimaging studies on healthy populations have confirmed a similar hierarchical organization, with longer INT in frontal and parietal compared to sensory areas (Hasson et al., 2008; Honey et al., 2012). This has been argued to form the basis of a functional hierarchy in the brain (Kiebel, Daunizeau and Friston, 2008; Lerner et al., 2011) that enables sensory areas to register fast environmental changes (Salinas et al., 2000) and cognitive areas to integrate and analyse sensory input (Wang, 2002). This functional hierarchy has a practical relevance for both localized and distributed neural activity, as shown by Ito, Hearne and Cole (2020). These authors showed that regions with faster INT during resting state displayed strong activations and decreased functional connectivity during task states. Additionally, Fallon et al. (2020) further showed that increased INT correlated with increased structural connectivity, thus expanding on the practical implications of assessing cortical temporal dynamics. This intrinsic hierarchical functional organization of brain activity and its alterations can improve the state of our current knowledge on how and where information processing breaks down in the healthy, but mostly in the dysfunctional brain.

This functional hierarchy has been shown to be altered in SZ. For example, Wengler et al. (2020) found reduced INT at the whole brain level in SZ compared to HC, and showed that INT reduction in auditory areas was modulated by hallucination and delusion severity. We were therefore interested to see how well this pattern is replicable across independent SZ samples and whether symptom severity plays a modulatory role. For this purpose, rsfMRI data has been analysed along the lines of Watanabe, Rees and Masuda (2019). Replicability can be however dramatically compromised if false positives are not controlled for. One source of spurious results in resting-state fMRI analyses are head motion artefacts, which are particularly frequent in clinical populations (e.g. Power et al., 2012; Van Dijk, Sabuncu and Buckner, 2012). Since we analysed data collected from SZ patients, this was a concern which we sought to address, therefore we analysed the INT group-differences both before and after eliminating framewise displacement outliers in all three SZ samples.

## Methods

### Participants

Three independently collected, age and sex-balanced samples were used in the present study; (1) an in-house all-male dataset collected at the Centre for Cognitive Neurosciences in Salzburg, Austria; (2) an open-source data-set from the Center for Biomedical Research Excellence (COBRE; Calhoun et al., 2011; Hanlon et al., 2011; Mayer et al., 2013; Stephen et al., 2013; available at http://fcon_1000.projects.nitrc.org/indi/retro/cobre.html); (3) an open-source data-set from the UCLA Consortium for Neuropsychiatric Phenomics LA5c Study (UCLANP; available at https://www.openfmri.org/dataset/ds000030/). Sample demographics and phenotypic information for all three samples are provided in Table 1.

**Table 1.**
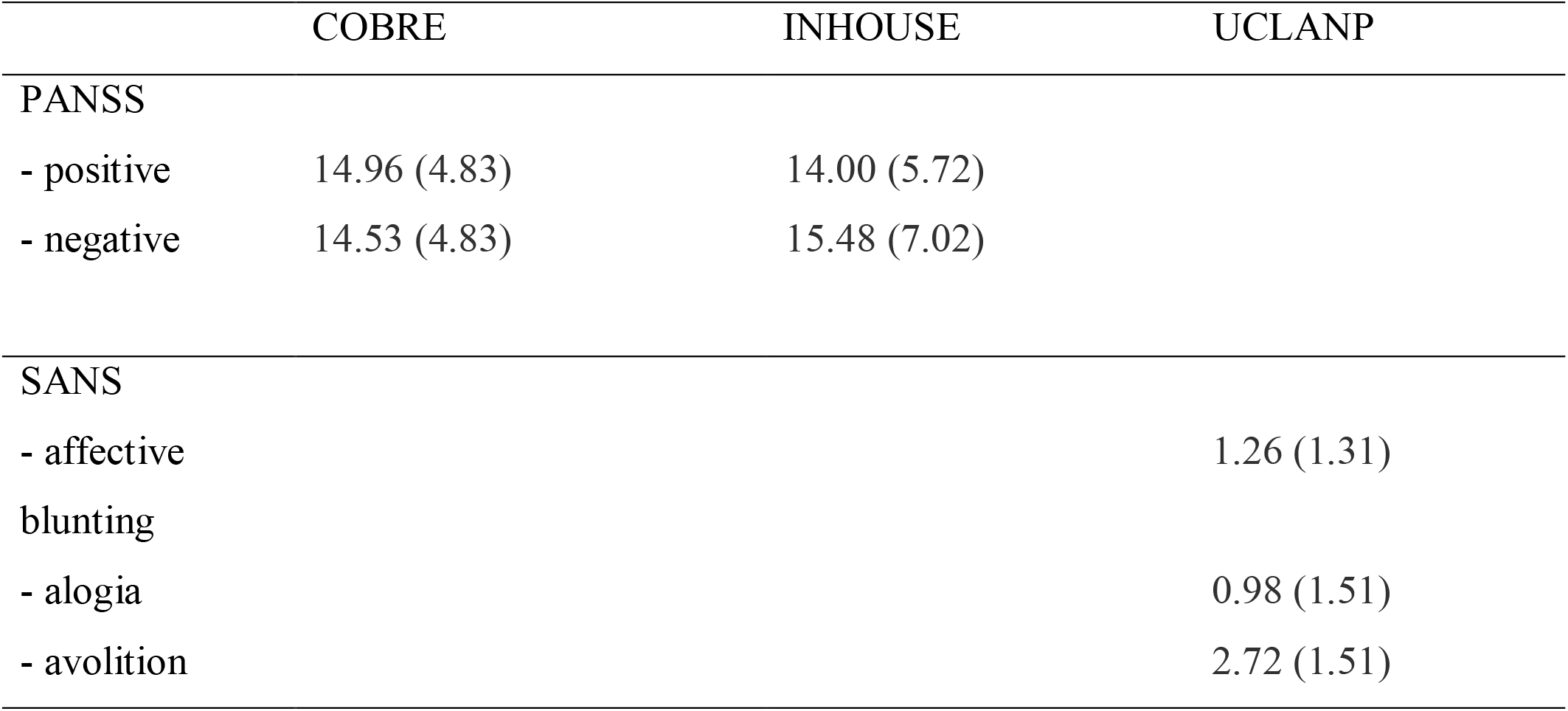
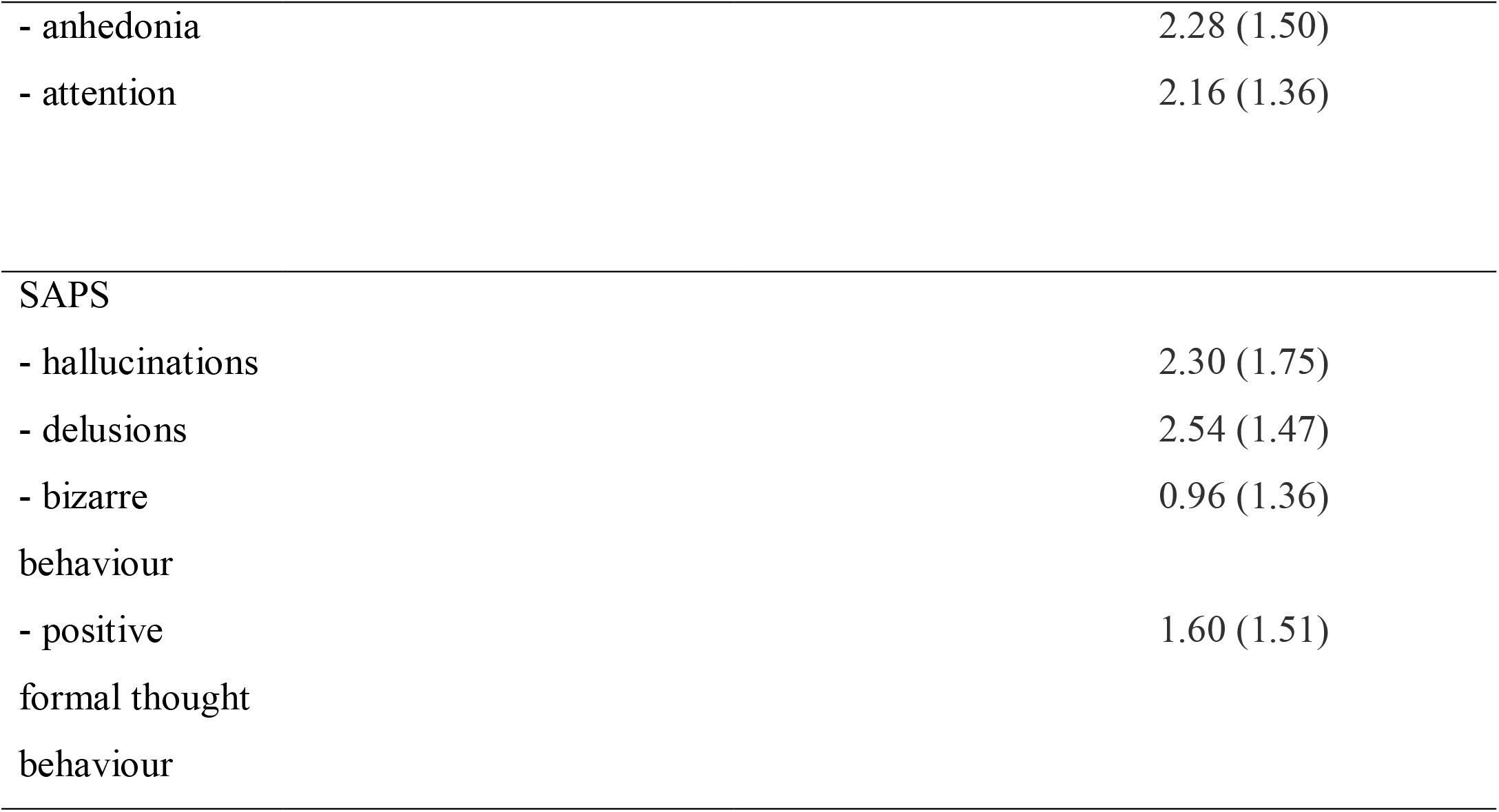
Mean and standard deviations of symptom severity in all three patient samples.

The in-house sample consisted of 25 all-male patients (Age mean and sd: 26.26 (4.83)) recruited at the Department of Psychiatry, Psychotherapy and Psychosomatics at the Christian-Doppler Medical Centre in Salzburg, Austria who had received a formal ICD-10 diagnosis in the schizophrenia spectrum group (F20) or the schizoaffective disorders spectrum group (F25). At the time of scanning, patients were medicated and clinically stable, with mild symptom severity, as assessed with PANSS (Kay, Fiszbein and Opler, 1987). Two of the patients did not complete the PANSS assessment but did complete the resting state scanning session. Thirty-one age and education matched control participants (Age mean and sd: 25.10 (4.33)) were recruited and screened for mental and physical health (*via* a standardized anamnesis procedure) and were excluded if they reported a history of mental or neurological disorder or a family history of psychiatric disorders. More details about the recruitment and assessment of the participants included in the present study can be found in Kronbichler et al. (2018). Functional imaging data were acquired on a Siemens Magnetom Trio 3T scanner (Siemens AG, Erlangen, Germany) using a 32-channel head coil. Functional images were acquired with a T2*-weighted gradient echo EPI sequence (TR 2,250 ms, TE 30 ms, matrix 64 mm × 64 mm, FOV 192 mm, flip angle 70°). Thirty-six slices with a slice thickness of 3 mm and a slice gap of 0.3 mm were acquired within the TR. Scanning was completed over two sessions with 321 scans per session. Finally, a gradient echo field map (TR 488 ms, TE 1 = 4.49 ms, TE 2 = 6.95 ms) and a high-resolution (1 mm × 1 mm × 1 mm) structural scan with a T1-weighted MPRAGE sequence were also acquired.

Seventy-two SZ (58 males; Age mean and sd: 38.17 (13.98)) and seventy-four HC (51 males; Age mean and sd: 35.82 (11.58)) from the COBRE open source dataset were included in this study. All participants were screened for a history of neurological disorders, mental retardation, and severe head trauma with more than 5 minutes loss of consciousness, substance abuse or dependence within the last 12 months. Clinical diagnosis was established using the Structured Clinical Interview used for DSM Disorders (SCID). A multi-echo MPRAGE (MEMPR) sequence was ran with the following parameters: TR/TE/TI = 2530/[1.64, 3.5, 5.36, 7.22, 9.08]/900 ms, flip angle = 7°, FOV = 256×256 mm, Slab thickness = 176 mm, Matrix = 256×256×176, Voxel size =1×1×1 mm, Number of echos = 5, Pixel bandwidth =650 Hz, Total scan time = 6 min. Resting state data was collected with single-shot full k-space echo-planar imaging (EPI) with ramp sampling correction using the anterior-to-posterior commissural line as a reference (TR: 2 s, TE: 29 ms, matrix size: 64×64, 32 slices, voxel size: 3×3×4 mm3).

Fifty SZ (38 males; Age mean and sd: 36.46 (8.88)) and sixty-three (44 males; Age mean and sd: 33.73 (9.1)) HC were included from the UCLA Consortium for Neuropsychiatric Phenomics LA5c Study. Participants were screened for neurological disease and major mental illness, history of head injury with loss of consciousness, use of psychoactive medications, and substance dependence within 6 months prior to testing. Self-reported history of psychopathology was assessed with the SCID-IV (First, Spitzer, Gibbon, & Williams, 1995). Urinalysis was used to screen for drugs of abuse (cannabis, amphetamine, opioids, cocaine, benzodiazepines) on the day of testing and participants were excluded if their results were positive. Neuroimaging data were acquired on a 3T Siemens Trio scanner. Functional MRI data were collected with a T2*-weighted echo planar imaging (EPI) sequence with the following parameters: slice thickness = 4mm, 34 slices, TR=2s, TE=30ms, flip angle=90°, matrix=64 × 64, FOV=192mm. A T1-weighted high-resolution anatomical scan (MPRAGE) was collected with the following parameter: slice thickness = 1mm, 176 slices, TR=1.9s, TE=2.26ms, matrix=256 x 256, FOV=250mm. The eyes open resting state fMRI session lasted for 304 seconds.

Symptom severity was assessed using The Scale for the Assessment of Negative Symptoms (SANS; Andreasen, 1984) and the Scale for the Assessment of Positive Symptoms (SAPS; Andreasen, 1984) for the UCLANP sample, and the Positive and negative symptom scale (PANSS; Kay et al., 1987) for the COBRE and the inhouse samples. Means and standard deviations of symptom severity measures for all three patient samples are given in Table 1.

### Neuroimaging data pre-processing

The fMRI data was pre-processed and analysed using SPM12 (Wellcome Trust Centre for Neuroimaging, London, UK; code available at: https://github.com/spm/spm12), while all the other statistical analyses were performed in R 5.2 (R Core Team, 2018). Functional scans were realigned, de-spiked, unwarped, corrected for geometric distortions, and slice time corrected. They were also normalized to MNI space and co-registered to the corresponding skull-stripped structural images, and afterwards resampled to 3 mm × 3 mm × 3 mm voxels and smoothed with a 6 mm FWHM Gaussian kernel. Motion correction was performed using ICA-AROMA (Pruim et al., 2015; http://fsl.fmrib.ox.ac.uk/fsl/fslwiki/OtherSoftware), and the resulting non-aggressively corrected resting state time series were used for computing the INT.

### Analysis

Following the INT analysis of Watanabe et al. (2019), the autocorrelation function was calculated for each voxel at incremental time lags until the autocorrelation function value became negative for the first time. The positive autocorrelation values were then summed up. The resulting sum was then multiplied by the repetition time (TR) to account for temporal resolution differences between the three samples. An index of the INT was thus obtained.

Next, using the COBRE dataset, we identified group differences in INT duration between HC and SZ via voxel-wise analysis. We isolated five clusters which we then used as masks to extract INT duration from those specific locations in HC and SZ from the in-house and the UCLANP datasets. This extraction was performed using the REX toolbox. Individual INT values were then exported and subsequently analysed using the R software.

As previous concerns have been raised regarding the possibility that head motion artefacts can cause false positive results, we re-analysed INT duration group differences after eliminating motion artefacts outliers. These artefacts (i.e. framewise displacement parameters; FD) were identified using the FSL library (Jenkinson et al., 2012). Group differences in FD between HC and SZ were minimized in two steps. First, the SZ with the largest FD values were gradually eliminated until the FD group difference was no longer significant (i.e. *p* > .05). Then, the HC with the smallest FD values were gradually eliminated until the mean FD values of the two groups were as similar as possible (*p* ≈ 1).

## Results

First, we ensured that sex and age did not differ significantly between SZ and HC in either of the three samples. For the in-house data set, as it was all-male, we only checked that the two groups were age-balanced (Mean (SD) of SZ = 26.26 (4.83); Mean (SD) of HC = 25.1 (4.33); t (44.494) = -0.91523, *p* = .365). The COBRE sample was equally balanced for both sex (χ^2^(1) = 2.033, *p* = .134) and age (Mean (SD) of SZ = 38.2 (13.9); Mean (SD) of HC = 35.8 (11.6); t (138.07) =-1.105, *p* = .271). The UCLANP sample was equally sex (χ^2^(1) = 0.267, *p* = .61) and age (Mean (SD) of SZ = 36.46 (8.88); Mean (SD) of HC = 33.73 (9.1); t (106.31) = -1.606, *p* = .111) balanced.

### Voxel-wise exploratory results

Exploratory mass-univariate t tests were first performed in COBRE, to compare the INT index between HC and SZ. First, the expected pattern of increased INT (Lerner et al., 2011) in frontal and parietal areas, and decreased INT in sensory areas was also confirmed in both HC and SZ (see Figure 1 below, top left and right panels). A pattern of increased INT in HC compared to SZ was observed in bilateral postcentral gyrus and occipital areas (see Figure 1 bottom left panel and table 1 below). Very few areas displayed increased INT in SZ compared to HC, in the supramarginal and inferior frontal gyrus (see Figure 1 below, bottom right panel).

**Figure 1.**
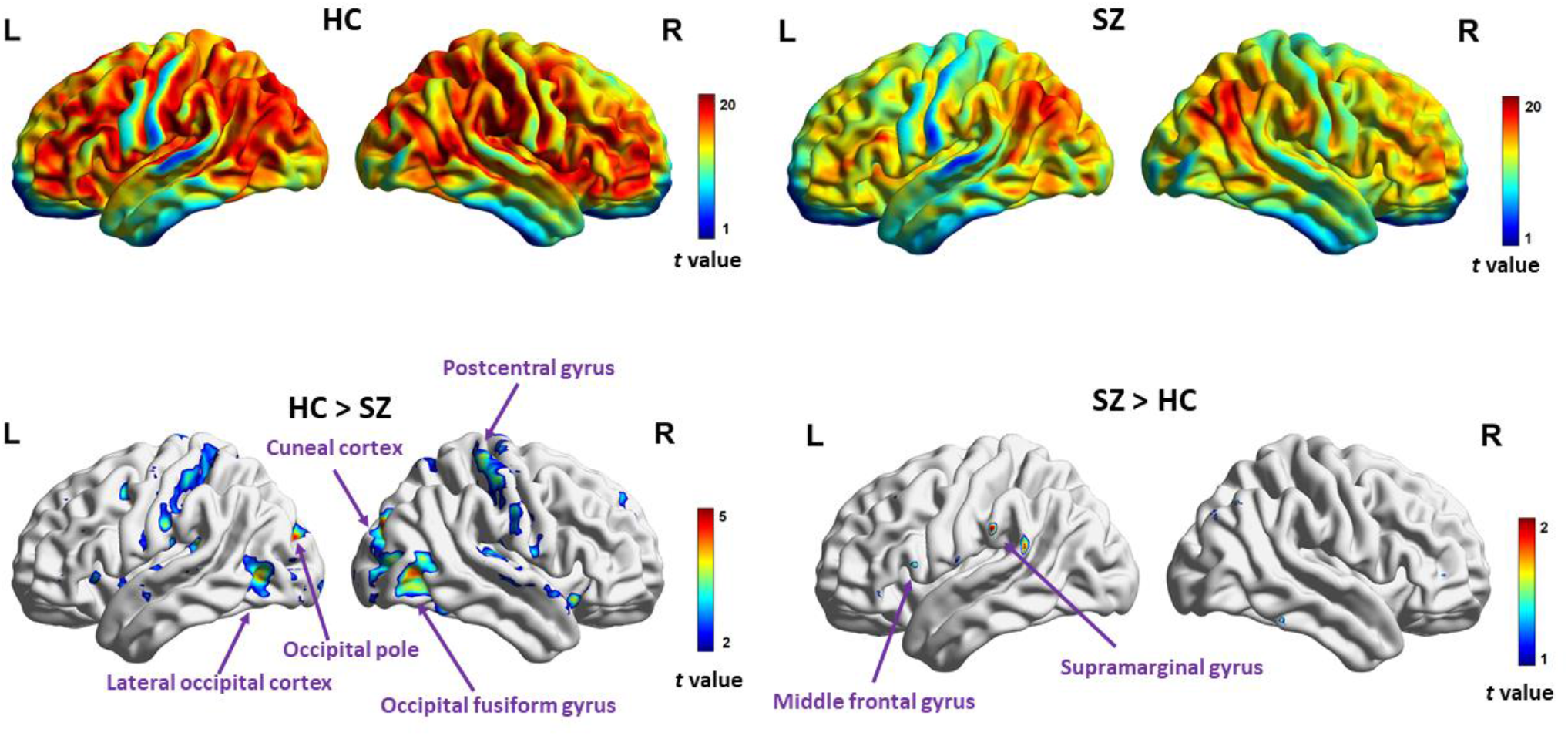
Mass-univariate exploratory analysis (*p* uncorr. < .001) of INT within and between SZ and HC of the COBRE dataset. In the top left and right, we observe the expected pattern of increased INT in frontal and parietal areas in both groups. The lower left panel depicts the areas that show longer INT in HC compared to SZ, namely: the right postcentral gyrus, the right occipital fusiform gyrus and right cuneal cortex, the left occipital pole and the left lateral occipital cortex. The lower right panel depicts the areas that show longer INT in SZ compared to HC, namely the left supramarginal and middle frontal gyrus.

### Region of interest (ROI) identification

Based on the mass univariate results from the COBRE dataset, prior to FD outlier elimination, we selected the significant clusters and thus extracted the five ROIs (see table 2 below) which were used in the subsequent cluster-level analyses. In order to avoid double-dipping, we excluded the COBRE dataset from the first set of ROI level group comparisons (i.e, prior to FD outlier elimination). We included it again when we re-ran our analyses post FD outlier elimination.

**Table 2.** Voxel-wise INT group differences.

| Anatomical label | Abbreviation | MNI coordinates |  |  | Cluster size | T value (FDR corr.) |
| --- | --- | --- | --- | --- | --- | --- |
| HC > SZ |  | X | Y | Z |  |  |
| Right occipital fusiform gyrus | rOFG | 27 | -88 | -14 | 122 | 4.34 |
| Left superior occipital gyrus | lSOG | -15 | -91 | 19 | 136 | 5.39 |
| Right superior occipital gyrus | rSOG | 18 | -79 | 25 | 452 | 5.04 |
| Left lateral occipital cortex | lLOC | -42 | -70 | 1 | 87 | 4.19 |
| Right postcentral gyrus | rPG | 42 | -31 | 64 | 61 | 4.61 |

### Group differences in INT duration between HC and SZ

Following ROI identification, we proceeded to analyse group differences in INT duration between HC and SZ within each of the five ROIs, for both the in-house and the UCLANP samples. Welch two-samples t tests were used for this purpose, as implemented in the R software. A consistent pattern of reduced INT in SZ compared to HC was found in all ROIs, and replicated in both samples. Within the in-house dataset, significantly increased INT durations in HC compared to SZ were found in the rSOG (Mean (SD) of INT in HC = 2.66 (0.61), Mean (SD) of INT in SZ = 2.22 (0.55), t (54.19) = 2.86, *p* Bonf. = .03, Hedge’s g = 0.74) and the rPG (Mean (SD) of INT in HC = 2.42 (0.67), Mean (SD) of INT in SZ = 2 (0.43), t (54.74) = 2.87, *p* Bonf. = .03, Hedge’s g = 0.71). In UCLANP, significantly longer INT were found in HC compared to SZ in the rOFG (Mean (SD) of INT in HC = 0.84 (0.42), Mean (SD) of INT in SZ = 0.67 (0.34), t (110.92) = 2.37, *p* Bonf. = .05, Hedge’s g = 0.44), the rSOG (Mean (SD) of INT in HC = 0. 7(0.39), Mean (SD) of INT in SZ = 0.52 (0.27), t (108.46) = 2.95, *p* Bonf. = .05, Hedge’s g = 0.53) and the rPG (Mean (SD) of INT in HC = 0.99 (0.59), Mean (SD) of INT in SZ = 0.76 (0.36), t (105.34) = 2.54, *p* Bonf. = .03, Hedge’s g = 0.46). These results are illustrated in Figure 2 below and summarized in Table 3 in the Supplement.

**Table 3.** INT group differences per sample and cluster, *before FD outlier elimination*. Both uncorrected and Bonferroni corrected p values are given. Due to the HC and SZ sample sizes not being identical, Hedge’s g was preferred as an effect size estimator. The HC > SZ comparison was assessed using one-sided Welch two-samples t tests.

|  | Mean (SD) INT |  | HC > SZ |  |  |  |  |
| --- | --- | --- | --- | --- | --- | --- | --- |
|  | HC | SZ | t | df | *P*<br>(*uncorr*) | *p*<br>(*Bonf. Corr*) | *Hedge's g* |
| *INHOUSE* |  |  |  |  |  |  |  |
| Cluster 1 | 2.64 (0.61) | 2.24 (0.54) | 2.63 | 54.43 | .011 | .055 | 0.68 |
| Cluster 2 | 2.51 (0.84) | 1.98 (0.73) | 2.55 | 54.8 | .014 | .07 | 0.66 |
| Cluster 3 | 2.66 (0.61) | 2.22 (0.55) | 2.86 | 54.19 | .006 | .03 | 0.74 |
| Cluster 4 | 2.61 (0.77) | 2.34 (0.52) | 2.23 | 55.37 | .03 | .15 | 0.55 |
| Cluster 5 | 2.42 (0.67) | 2.0 (0.43) | 2.87 | 54.74 | .006 | .03 | 0.71 |
| *UCLANP* |  |  |  |  |  |  |  |
| Cluster 1 | 0.84 (0.42) | 0.67 (0.34) | 2.37 | 110.92 | .01 | .05 | 0.44 |
| Cluster 2 | 0.86 (0.44) | 0.75 (0.39) | 1.42 | 110.08 | .08 | .4 | 0.26 |
| Cluster 3 | 0.7 (0.39) | 0.52 (0.27) | 2.95 | 108.46 | .01 | .05 | 0.53 |
| Cluster 4 | 0.81 (0.47) | 0.77 (0.44) | 0.51 | 107.85 | .31 | 1 | 0.1 |
| Cluster 5 | 0.99 (0.59) | 0.76 (0.36) | 2.54 | 105.34 | .006 | .03 | 0.46 |

**Figure 2.**
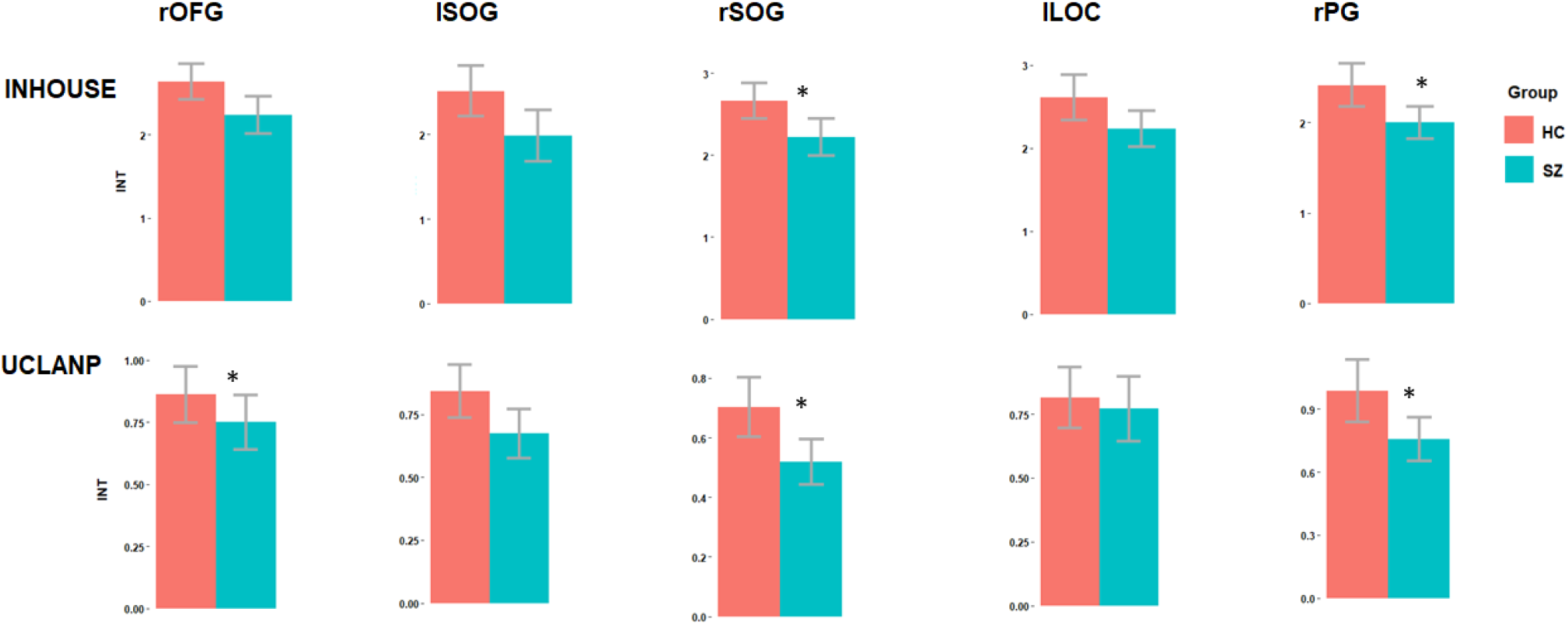
Group differences in intrinsic neural time scales (INT) per sample and cluster, *prior to FD outlier elimination*. Bonferroni-corrected significant differences are marked. Red colour represents HC, while blue represents SZ.

### Relationship between symptom severity and INT

We also explored the relationship between symptom severity and INT of the five clusters, in the three patient samples. We found no significant relationship between INT duration in any of the five clusters and symptom severity in our inhouse sample (r < 0.21, *p* > .31) and in COBRE (r < 0.24, *p* > .22). In the UCLANP samples, we found a significant negative correlation between the INT duration of the Right occipital fusiform gyrus and positive formal thought disorder (r = -0.3, *p* = .03).

### Elimination of outliers with extreme head motion artefacts

Due to concerns regarding the false positive rate which might be driven upwards by motion artefacts, we re-analysed the INT group differences after FD outlier elimination. The previously observed pattern of increased INT in HC compared to SZ is preserved across all clusters and samples. In COBRE, significantly longer INT in HC compared to SZ were found in all five clusters: rOFG (Mean (SD) of INT in HC = 1.56 (0.57), Mean (SD) of INT in SZ = 1.16 (0.49), t (111.6) = 4.1, *p* Bonf. < .001, Hedge’s g = 0.74), lSOG (Mean (SD) of INT in HC = 1.53 (0.64), Mean (SD) of INT in SZ = 1.17 (0.55), t (111.6) = 3.33, *p* Bonf. = .005, Hedge’s g = 0.61), rSOG (Mean (SD) of INT in HC = 1.41 (0.54), Mean (SD) of INT in SZ = 1.01 (0.46), t (111.27) = 4.32, *p* Bonf. < .001, Hedge’s g = 0.8), lLOC (Mean (SD) of INT in HC = 1.62 (0.68), Mean (SD) of INT in SZ = 1.19 (0.54), t (108.79) = 3.81, *p* Bonf. < .001, Hedge’s g = 0.71) and rPG (Mean (SD) of INT in HC = 1.63 (0.69), Mean (SD) of INT in SZ = 1.14 (0.53), t (107.5) = 4.31, *p* Bonf. < .001, Hedge’s g = 0.8). In the inhouse sample, significantly longer INT in HC compared to SZ were found in rSOG (Mean (SD) of INT in HC = 2.59 (0.43), Mean (SD) of INT in SZ = 2.1 (0.44), t (26.29) = 2.14, *p* Bonf. = .005, Hedge’s g = 1.15) and rPG (Mean (SD) of INT in HC = 2.45 (0.65), Mean (SD) of INT in SZ = 1.9 (0.45), t (28.23) = 2.89, *p* Bonf. = .04, Hedge’s g = 0.96). In UCLANP, significantly longer INT in HC compared to SZ were found in rOFG (Mean (SD) of INT in HC = 0.84 (0.42), Mean (SD) of INT in SZ = 0.67 (0.34), t (107) = 2.32, *p* Bonf. = .05, Hedge’s g = 0.44), rSOG (Mean (SD) of INT in HC = 0.7 (0.4), Mean (SD) of INT in SZ = 0.51 (0.27), t (103.21) = 2.88, *p* Bonf. = .01, Hedge’s g = 0.53) and rPG (Mean (SD) of INT in HC = 0.97 (0.58), Mean (SD) of INT in SZ = 0.75 (0.37), t (101.34) = 2.39, *p* Bonf. = .05, Hedge’s g = 0.44). These results are illustrated in Figure 3 below and summarized in Table 4 in the Supplement.

**Table 4.**
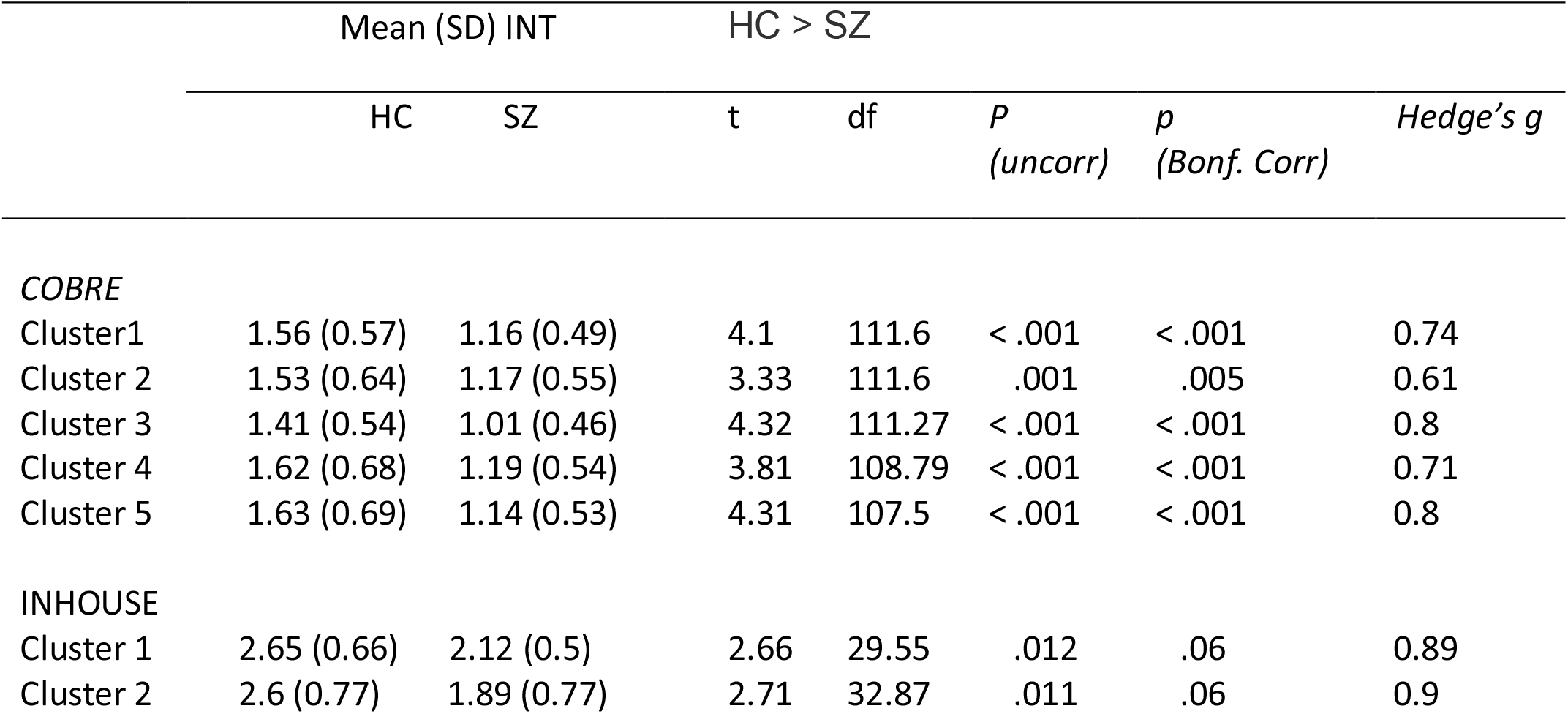
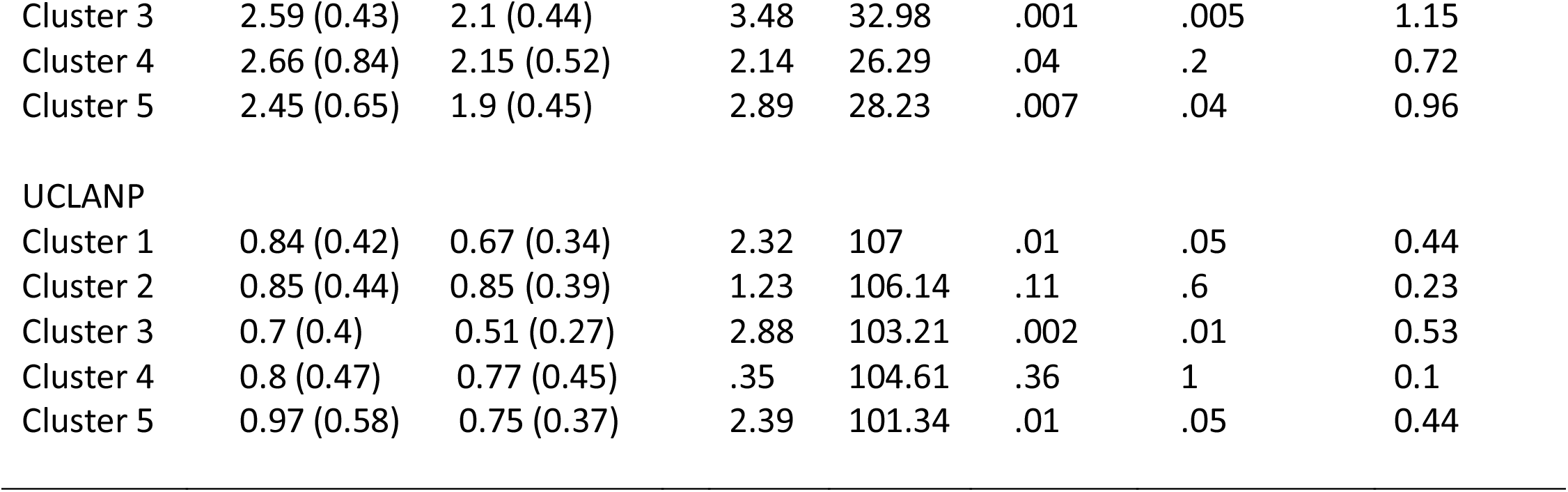
INT group differences per sample and cluster, *after FD outlier elimination*. Both uncorrected and Bonferroni corrected p values are given. Due to the HC and SZ sample sizes not being identical, Hedge’s g was preferred as an effect size estimator. The HC > SZ comparison was assessed using one-sided Welch two-samples t tests.

|  | Mean (SD) INT |  | HC > SZ |  |  |  |  |
| --- | --- | --- | --- | --- | --- | --- | --- |
|  | HC | SZ | t | df | *P*<br>(*uncorr*) | *p*<br>(*Bonf. Corr*) | *Hedge's g* |
| *COBRE* |  |  |  |  |  |  |  |
| Cluster1 | 1.56 (0.57) | 1.16 (0.49) | 4.1 | 111.6 | < .001 | < .001 | 0.74 |
| Cluster 2 | 1.53 (0.64) | 1.17 (0.55) | 3.33 | 111.6 | .001 | .005 | 0.61 |
| Cluster 3 | 1.41 (0.54) | 1.01 (0.46) | 4.32 | 111.27 | < .001 | < .001 | 0.8 |
| Cluster 4 | 1.62 (0.68) | 1.19 (0.54) | 3.81 | 108.79 | < .001 | < .001 | 0.71 |
| Cluster 5 | 1.63 (0.69) | 1.14 (0.53) | 4.31 | 107.5 | < .001 | < .001 | 0.8 |
| *INHOUSE* |  |  |  |  |  |  |  |
| Cluster 1 | 2.65 (0.66) | 2.12 (0.5) | 2.66 | 29.55 | .012 | .06 | 0.89 |
| Cluster 2 | 2.6 (0.77) | 1.89 (0.77) | 2.71 | 32.87 | .011 | .06 | 0.9 |
| Cluster 3 | 2.59 (0.43) | 2.1 (0.44) | 3.48 | 32.98 | .001 | .005 | 1.15 |
| Cluster 4 | 2.66 (0.84) | 2.15 (0.52) | 2.14 | 26.29 | .04 | .2 | 0.72 |
| Cluster 5 | 2.45 (0.65) | 1.9 (0.45) | 2.89 | 28.23 | .007 | .04 | 0.96 |
| UCLANP |  |  |  |  |  |  |  |
| Cluster 1 | 0.84 (0.42) | 0.67 (0.34) | 2.32 | 107 | .01 | .05 | 0.44 |
| Cluster 2 | 0.85 (0.44) | 0.85 (0.39) | 1.23 | 106.14 | .11 | .6 | 0.23 |
| Cluster 3 | 0.7 (0.4) | 0.51 (0.27) | 2.88 | 103.21 | .002 | .01 | 0.53 |
| Cluster 4 | 0.8 (0.47) | 0.77 (0.45) | .35 | 104.61 | .36 | 1 | 0.1 |
| Cluster 5 | 0.97 (0.58) | 0.75 (0.37) | 2.39 | 101.34 | .01 | .05 | 0.44 |

**Figure 3.**
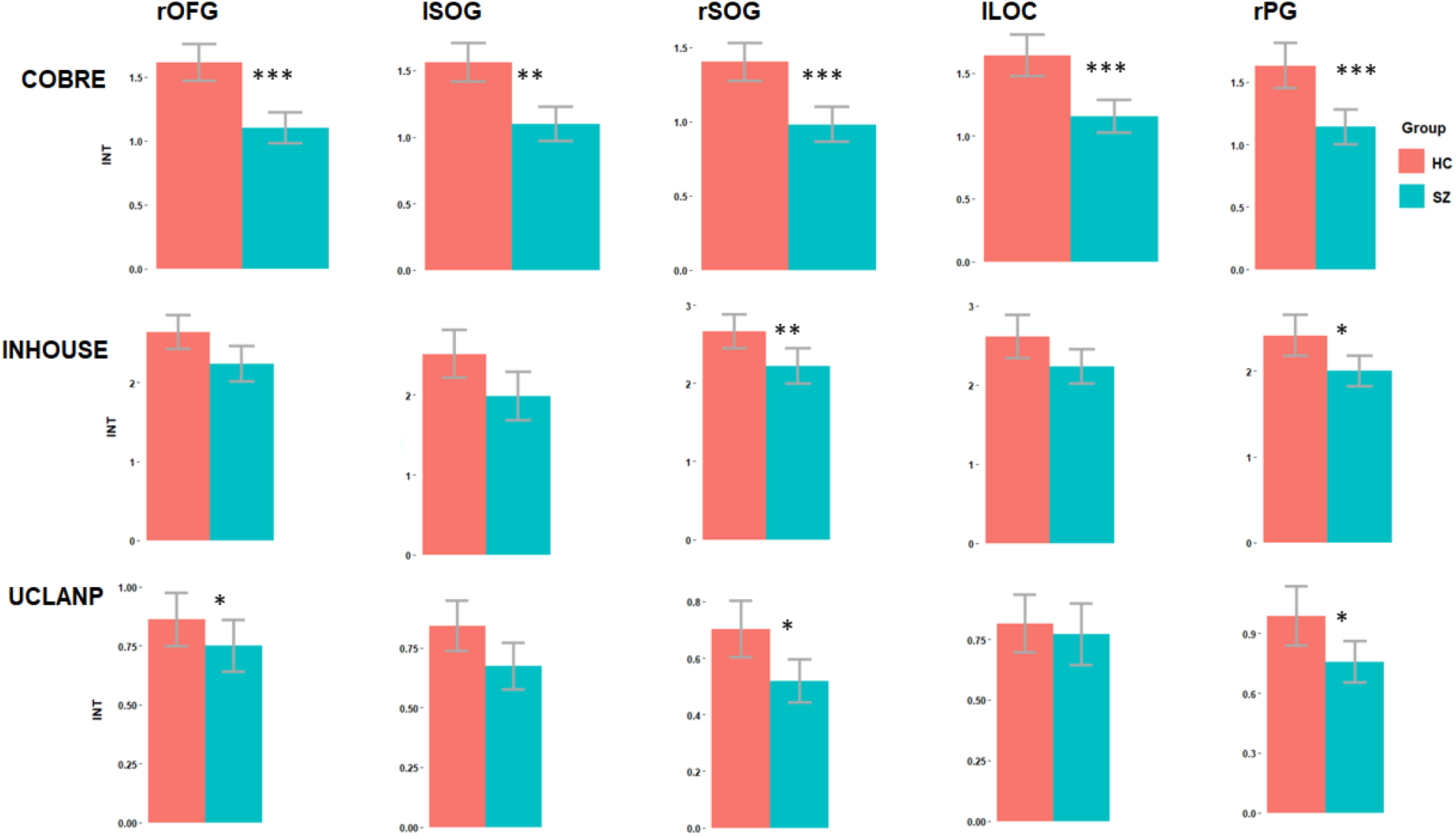
Group differences in intrinsic neural time scales (INT) per sample and cluster, *after FD outlier elimination*. Bonferroni-corrected significant differences are marked. Red colour represents HC, while blue represents SZ.

Finally, to ensure that INT group differences are not due to motion artefacts, we further explored the relationship between FD and INT. In COBRE and UCLANP there were no significant correlations between INT duration and FD, neither before nor after FD outlier elimination (see Figure 4 below). In the in-house SZ sample, there were three significant positive correlations between FD and INT duration in the rOFG (r = 0.43, *p* = .03), rSOG (r = 0.56, *p* = .004) and lLOC (r = 0.42, *p* = .04). However, this relationship became non-significant after FD outlier elimination. Thus, even if motion artefacts might have had an initial influence on group differences, we believe that this was removed in the second step, by eliminating extreme FD values.

**Figure 4.**
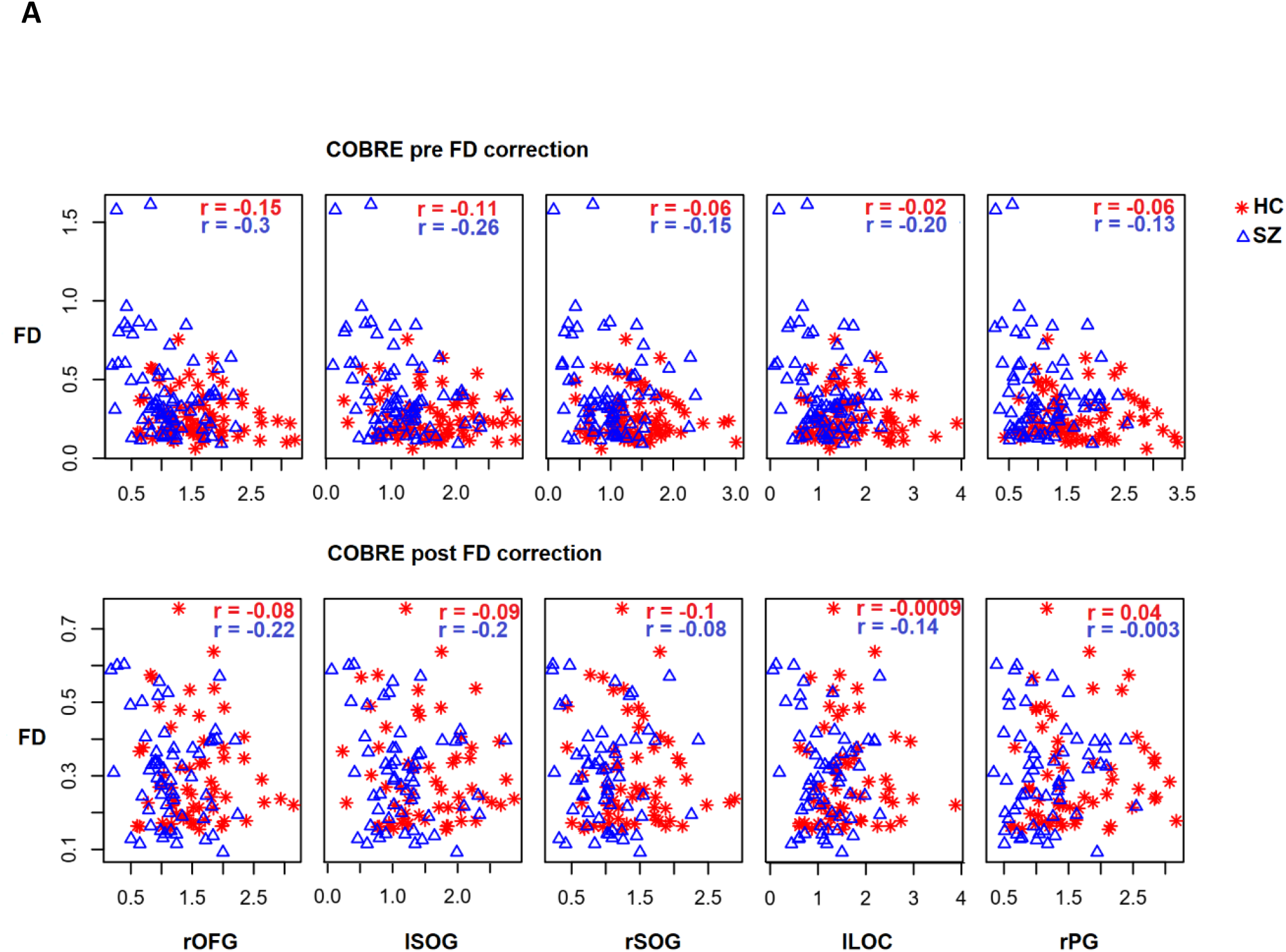
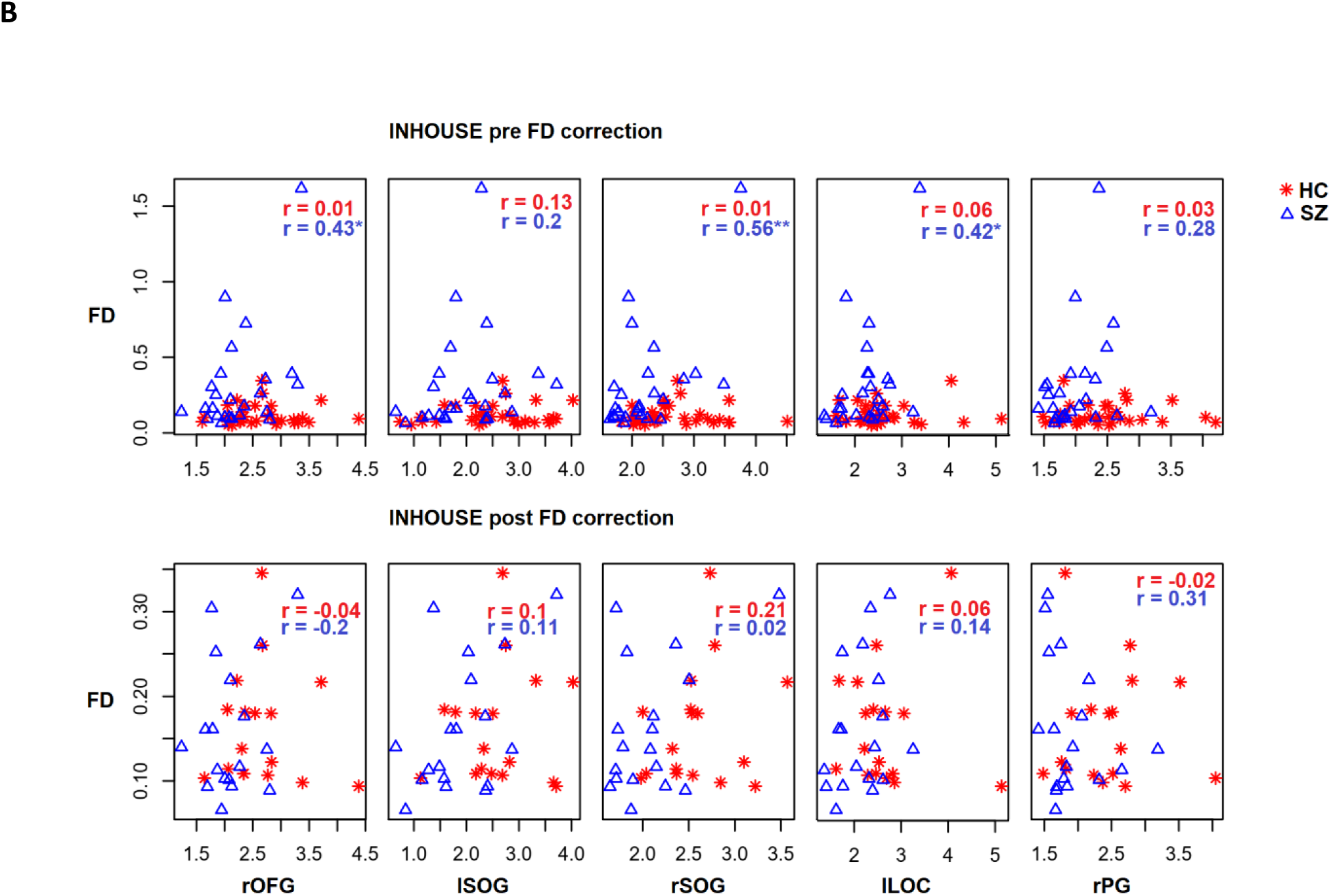
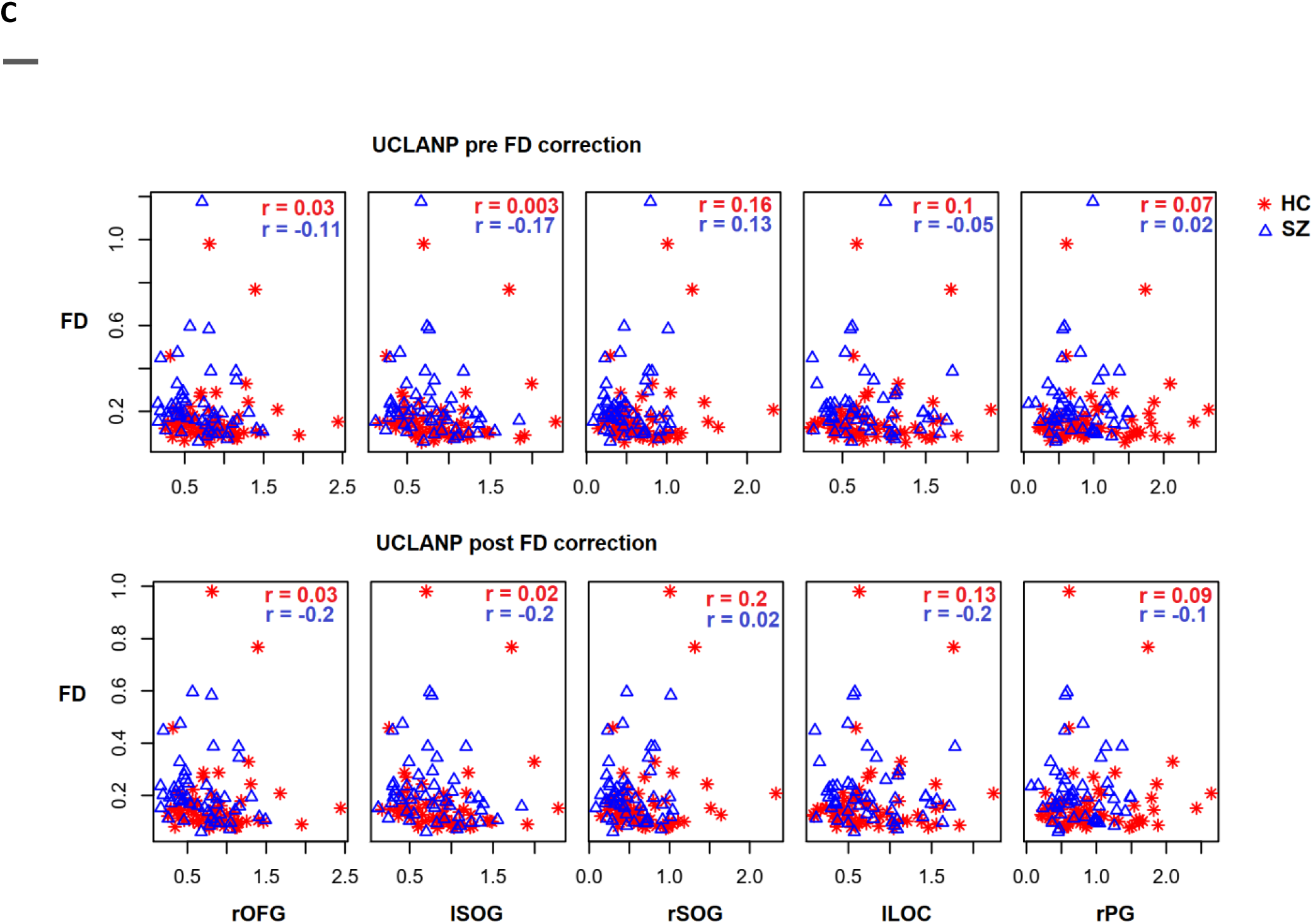
Correlations between framewise displacement (FD) and intrinsic neural timescales (INT), before and after FD outlier elimination, per sample and cluster; COBRE in panel A, INHOUSE in panel B, and UCLANP in panel C. Red star symbols represent correlations between FD and INT in HC, while blue triangle symbols represent correlations between FD and INT in SZ. Correlation coefficients are written in red for HC and blue for SZ. Significant correlations are marked by asterisks.

### Relationship between symptom severity and INT after FD outlier elimination

We analysed the relationship between symptom severity and INT of the five clusters, in the three patient samples, following FD outlier elimination. No significant correlations were found in the in-house sample (r < -0.41, *p* > .1). In the UCLANP sample, we found a significant negative correlation between the INT duration of the rOFG and positive formal thought disorder (r = -0.3, *p* = .03).

## Discussion

In this paper, we present a replication of intrinsic neural timescales (INT) patterns in three independent samples of patients with schizophrenia (SZ) and matched healthy controls (HC). Our main goal was to assess to which extent INT findings can be reliably replicated across independent samples. We believe this to be a crucial step, as it has been shown that different data acquisition settings constitute a major obstacle in achieving replicability (King et al. 2019). We pre-processed and analysed all three samples identically and we were able to show that the pattern of reduced INT in SZ compared to HC also generalizes well across independently acquired samples, thus being robust to differences in acquisition protocols.

Motion artefacts were another concern which we sought to address, as they tend to occur frequently in clinical samples and often lead to spurious results (e.g. Power et al., 2012; Van Dijk et al., 2012). We therefore assessed the INT patterns before and after FD outlier elimination, and our results were significant in both instances. Additionally, we also checked whether a relationship between FD and INT might have led to false positives. In most cases, this relationship was not significant, even before eliminating FD outliers. In the few cases where there was a positive and significant relationship between FD and INT, this effect disappeared once we eliminated FD outliers. The pattern of reduced INT in SZ compared to HC was however invariably preserved. We were therefore satisfied that the results we observed reflected real and replicable group differences and not spurious results driven by motion artefacts.

We also assessed the relationship between INT and symptom severity and found a negative significant correlation between INT in the rOFG and positive formal thought disorder in the UCLANP sample. In other words, it appears that increased severity of this symptom is associated with increased excitation/inhibition (EI) ratio (hence decreased INT) in the rOFG. Previously, Wengler et al. (2020) looked at the relationship between hallucination and delusion severity in SZ and INT of auditory and visual areas. While they found such an association for the auditory system, they did not find any for the visual system. This may be explained by the different approaches in parcellation and ROI identification between our study and that of Wengler et al. While Wengler et al. opted for anatomically-defined parcels, we opted for a data-driven functional ROI identification based on group differences in the COBRE dataset. We believe that the difference in acquisition parameters of the different datasets (i.e., the Human Connectome Project in Wengler et al., and the COBRE, UCLANP and inhouse datasets of our study) had little influence over our result differences, since we already showed that INT patterns generalize well across different datasets.

One mechanistic interpretation with respect to the reduced INT in SZ was previously offered by Wengler et al. (2020). Following a series of simulations, these authors suggest that a reduction in the excitation-inhibition (E/I) ratio could account for the global, brain-level INT reduction in SZ compared to HC. Computational studies have also previously linked long-range autocorrelation fluctuations to the E/I ratio (e.g. Deco et al., 2014). This mechanism appears to be supported by clinical findings as well, as similar E/I ratio imbalances have been found, e.g., in both ASD and SZ (Ford, Abu-Akel and Crewther, 2019). Given the overlap in sensory impairments between these two disorders, it is reasonable to propose that the INT patterns can be used as a trans-diagnostic biomarker bridging ASD and SZ (Foss-feig et al., 2017), and capturing underlying E/I imbalances in sensory areas.

An alternative explanatory mechanism that we propose for the reduced INT similarities between the two disorders could be sensory gating impairments, frequently documented in SZ and linked to thalamus and hypothalamus impairments (e.g. Edgar et al., 2005; Tregellas et al., 2007; 2009; Hazlett et al., 2008; Mayer et al., 2013; Çetin et all., 2014). Furthermore, Raut, Snyder and Raichle (2020) showed that the hierarchical cortical INT patterns are reflected within the thalamus, with INT increasing along a ventrolateral to dorsomedial axis, essentially from lower-to higher-order nuclei. Based on the evidence listed here, it appears that thalamic mediated sensory gating could lead to the observed INT group differences. However, further analyses linking the connectivity between the thalamus, hippocampus, frontal and sensory areas (Grunwald et al., 2003; Mayer et al., 2009) to the INT of sensory areas could provide the necessary evidence in favour of this proposed mechanism. As sensory integration alterations have been proposed to be a promising trans-diagnostic biomarker (Hornix et al., 2019), we argue that chronnectomic analyses offer the best approach, as these can reveal the dynamic fluctuations of thalamo-cortical and cortico-cortical connections, arguably capturing a more accurate representation of neural activity.

## Data Availability

The COBRE and UCLANP datasets are freely available online. The COBRE can be downloaded from http://fcon_1000.projects.nitrc.org/indi/retro/cobre.html. The UCLANP can be downloaded from https://exhibits.stanford.edu/data/catalog/mg599hw5271. The inhouse dataset can be obtained upon request.

## Supplement

